# MATHEMATICAL PREDICTIONS FOR COVID-19 AS A GLOBAL PANDEMIC

**DOI:** 10.1101/2020.03.19.20038794

**Authors:** Victor Alexander Okhuese

## Abstract

This study shows that the disease free equilibrium (***E***_**0**_) for COVID-19 coronavirus does not satisfy the criteria for a locally or globally asymptotic stability. This implies that as a pandemic as declared by WHO (2020) the COVID-19 coronavirus does not have a curative vaccine yet and precautionary measures are advised through quarantine and observatory procedures. Also, the Basic Reproductive number (*R*_0_ < 1) by Equation (33) shows that there is a chance of decline of secondary infections when the ratio between the incidence rate in the population and the total number of infected population quarantined with observatory procedure.

The effort to evaluate the disease equilibrium shows that unless there is a dedicated effort from government, decision makers and stakeholders, the world would hardly be reed of the COVID-19 coronavirus and further spread is eminent and the rate of infection will continue to increase despite the increased rate of recovery because of the absence of vaccine at the moment.

## 1. INTRODUCTION

A recent study by Nesteruk (2020) and Ming and Zhang (2020) focuses on the epidemic outbreak cased by COVID-19 coronavirus due to the global trend of the pandemic with its origin from mainland China. In his study, Nesteruk (2020) used the popular SIR (Susceptible-Infectious-Removed) model to obtain optimal values for the model parameters with the use of statistical approach and hence predicated the number of infected, susceptible and removed persons versus time. This model approach by Nesteruk (2020) has been a major breakthrough in modelling disease control as used by several authors (Ming and Zhang, 2020 and Oduwole and Kimbir (2018) among others). However, although there exist a global interest in knowing the rate of infection that will occur over time globally, it is of great interest to propose a mathematical model for the end in the spread and subsequent elimination of the virus. Hence, in this study we adopt solutions from Victor and Oduwole (2020) for a new deterministic endemic model (Susceptible – Exposed – Infectious – Removed – Undetectable – Susceptible: SEIRUS) originally developed for the control of the prevalence of HIV/AIDS in Africa.

The resulting equations are a system of coupled homogenous differential equations. Numerical experiments with relevant simulation showing how the variation of the reproductive number (R0) affect the number of infected individuals is carried out. Conscious effort through evaluating the new deterministic SEIRUS model is done to reduce the reproductive number (R0) to zero for a possible halt of the spread of the disease thereby leading to an endemic equilibrium to eradicate the disease in a later time in the future.

In summary, this study aims to develop and evaluate the new deterministic endemic SEIRUS compartmental model of the COVID-19 coronavirus dynamics which combines quarantine/observatory procedures and behavioral change/social distancing in the control and eradication of the disease in the most exposed sub-population.

The wide spread of the COVID-19 coronavirus and the lack or inefficiency of purposeful and result based intervention is a great call for other empirical and scientific interventions which seeks to review strategic models and recommendations of social and scientific research for disease control. Although previous studies have been tailored towards the epidemiology and the disease-free equilibrium where the reproductive number of the infectious population is at its barest minimum, this study seeks to study the evaluative impact of the endemic equilibrium of a new endemic deterministic model while taking into consideration that possibility of the recovered population being Undetectable and fit to be moved to the Susceptible compartment which will therefore imply a zero secondary infection of the disease globally.

Also, this research work will contribute significantly to recent research findings by Nesteruk (2020) and Ming and Zhang (2020) as well as inform government, non-government organizations as well as policy maker’s decision on sustainable policies to prevent the spread of the disease based on specific age groups of the active population.

## 2. THE MODEL VARIABLES AND PARAMETERS

The model variables and parameters for the investigation of the stability analysis of the equilibrium state for the new deterministic endemic model is given by;

| **Variable** | **Description** |
| --- | --- |
| $S(t)$ | Number of susceptible population at time $t$ |
| $E(t)$ | Number of exposed population at time $t$ |
| $I(t)$ | Number of infected population at time $t$ |
| $R(t)$ | Number of infected population quarantined and expecting recovery at time $t$ |
| $U(t)$ | Number of recovered adults satisfying undetectable criteria at time $t$ |

| **Parameter** | **Description** |
| --- | --- |
| $\mu$ | Natural death rate of the population |
| $\alpha_0$ | Maximum death rate due to COVID-19 coronavirus. ($\alpha \leq \alpha_0$) |
| $\alpha$ | Death rate of infected population due to COVID-19 coronavirus |
| $\varphi$ | Disease induced death rate of infected population not quarantined |
| $\varpi$ | Disease induced death rate of infected receiving quarantined |
| $T$ | Maximum lifespan after infection ($T \geq 14 \text{ days}$) |
| $k$ | Efficacy of Quarantine ($0 \leq k \leq 1$) |
| $\rho$ | Rate of recovery |
| $\beta$ | rate of transmission |
| $\sigma$ | Proportion of infected population in quarantine per unit time (Treatment rate) |
| $\pi$ | Proportion of population from susceptible to exposed/latent class |
| $\varepsilon$ | Proportion of removed population still being observed and being moved to susceptible class |
| $B(t)$ | Incidence rate or force of infection in the population |

### 2.1 MODEL ASSUMPTIONS

The following assumptions would help in the derivation of the model:

1. There is no emigration from the total population and there is no immigration into the population. A negligible proportion of individuals move in and out of the population at a given time.
2. Maturation (or maturity) is interpreted as the period between infection to the period of symptoms observation (days 1 to 14)
3. The susceptible population are first exposed to a latent class where they can infected or not.
4. Some infected individuals move to the removed class when they are quarantined for observatory procedures.
5. The recruitment from the *S*-class into the *E*-class is through contacts from population in the *I*-class to the *S-*class
6. The recruitment into the *R*-class from the *I*-class at a rate *σ*.
7. The recruitment into the *U*-class from the *R*-class depends on the effectiveness of the quarantine and observatory procedure at a rate *ρ*.
8. Death is implicit in the model and it occurs in all classes at constant rate *μ*. However, there is an additional death rate in the *I* and *R* classes due to infection for both juvenile and adult sub-population denoted by *φ* and *ϖ* respectively.

### 2.2 MODEL DESCRIPTION

This study uses the deterministic endemic model where a susceptible class is a class that is yet to be infected, but is open to infection as interactions with members of the *I*-class continuous. An infected individual is one who has contracted the coronavirus and is at some stage of infection. A removed individual is one that is confirmed to have the virus with its expected symptoms and is under quarantine while following relevant observatory procedures. A member of the undetectable class is one that has been removed and does not secrete the virus anymore and has been satisfied by the WHO standard to be in the undetectable class.

The following diagram describes the dynamic of SEIRUS framework, and will be useful in the formulation of model equations.

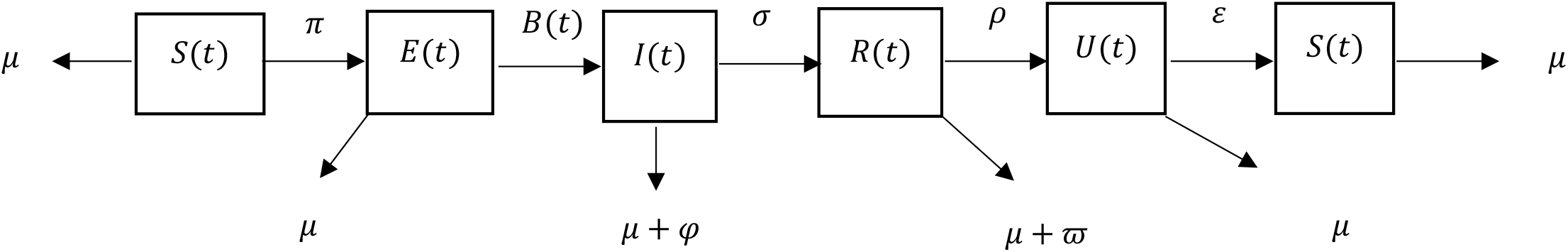

## 3. THE MODEL EQUATIONS

From the assumptions and the flow diagram above, the following model equations are derived.

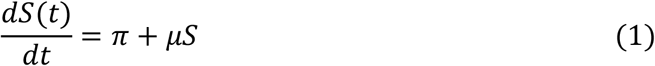

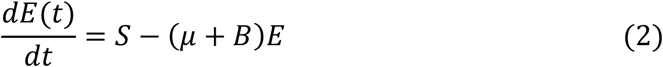

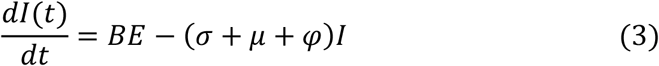

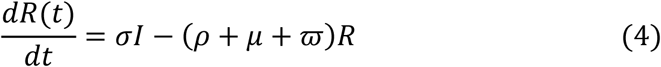

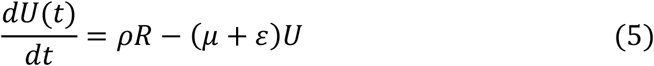

Such that

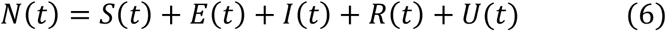

The incidence rate or force of infection at time *t* denoted by *B*(*t*) in the population is given as

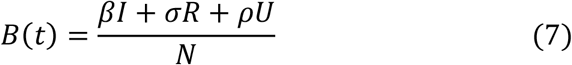

### 3.1 MODEL EQUATIONS IN PROPORTIONS

To simplify the model, we normalize the model by transforming the model equations into proportions. The derive model equations in proportion of infected juveniles and adults define prevalence of infection, which has biological meaning.

The model equations are transformed into proportions as follows;

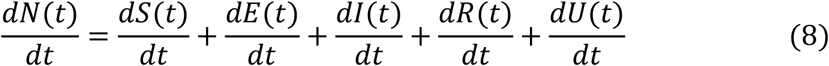

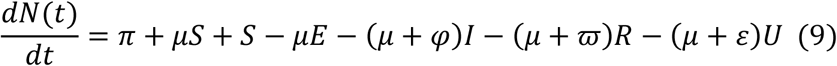

Let

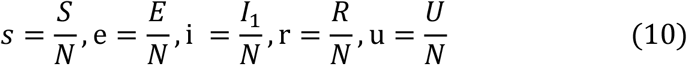

Then the normalized system is follows,

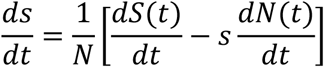

Substituting (1) and (9) and using (10)

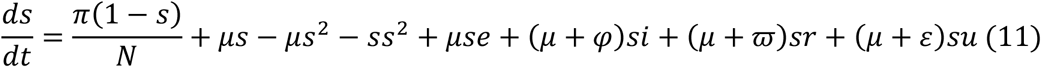

Similarly,

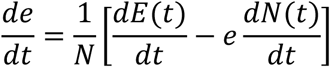

Substituting Equations (2) and (9) and using (10)

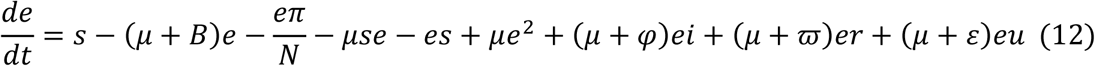

Next we have;

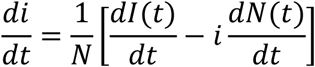

Substituting Equation (3) and (9) and using (10)

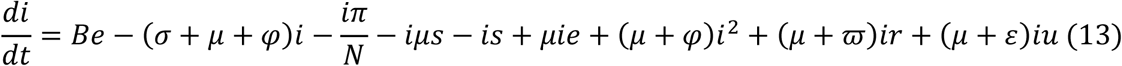

And

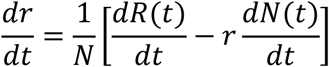

Substituting Equation (4) and (9) and using (10)

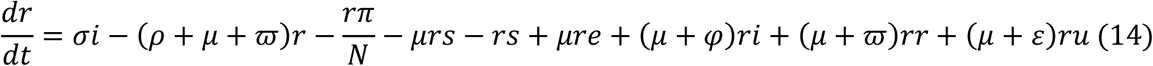

And finally;

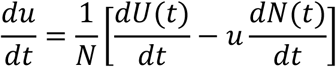

Substituting Equation (5) and (9) and using (10)

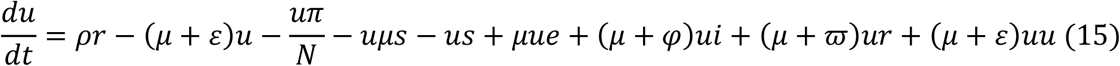

However,

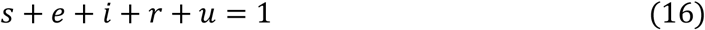

Equations (11) to (15) are the model equations in proportions, which define prevalence of infection.

### 3.2 EXISTENCE AND UNIQUENESS OF DISEASE FREE EQUILIBRIUM STATE (*E*_0_) OF THE SEIRUS MODEL

The disease-free equilibrium (DFE) state of the endemic SEIRUS model is obtained by setting the left hand sides of equations (11) – (15) to zero while setting the disease components *e* = *i* = *r* = *u* = 0 leading to equations (17) – (18) below

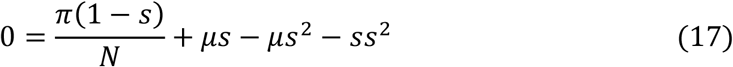

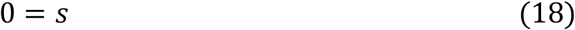

And substituting (18) into (17) we have;

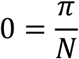

Then taking (17) where *s* = 0 or

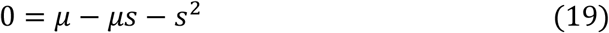

Simplifying further gives,

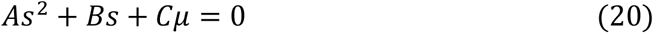

where *A* = 1, *B* = *μ* and *C* = −*μ*

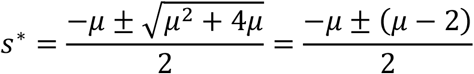

Therefore, the solution for the simultaneous equations (19) is given by

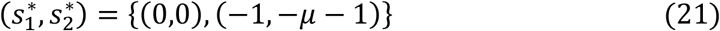

Ignoring the native values of 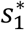 and 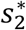 and other stringent conditions, there exist a unique trivial and disease-free equilibrium states at 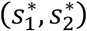 given by (0,0). The solution (21) satisfies equation (19) identically.

### 3.3 STABILITY ANALYSIS OF DISEASE FREE EQUILIBRIUM STATE (*E*_0_)

In order to study the behavior of the systems (11) – (15) around the disease-free equilibrium state *E*_0_ = [0,0,0,0,0], we resort to the linearized stability approach such that;

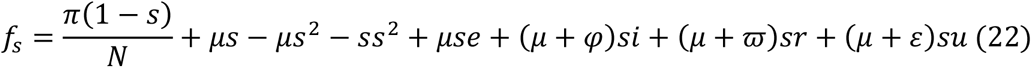

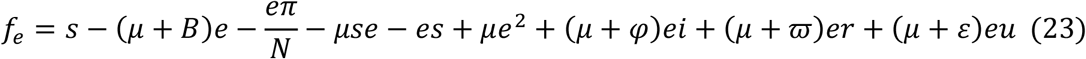

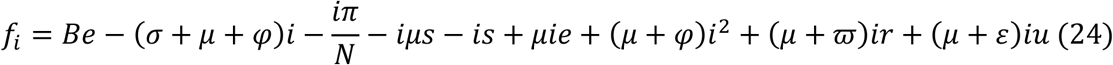

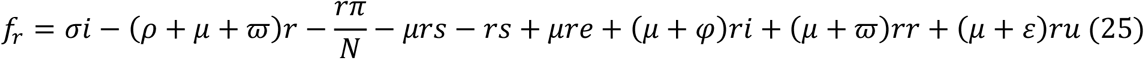

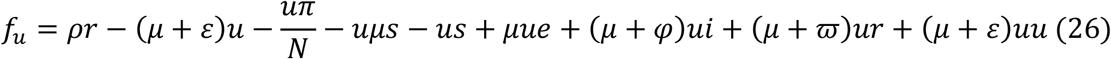

Therefore;

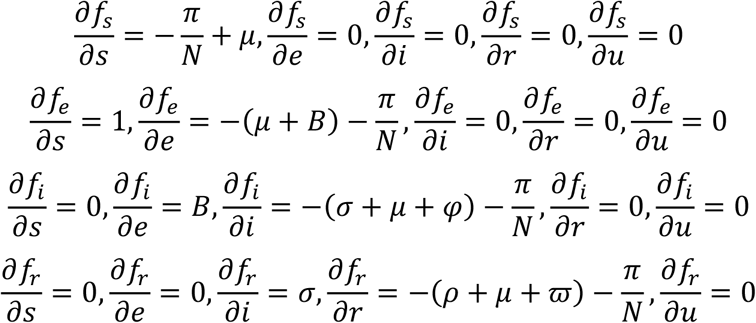

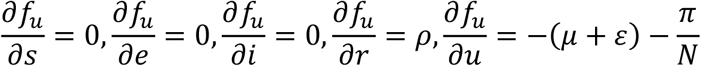

And the Jacobian 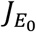 is given by

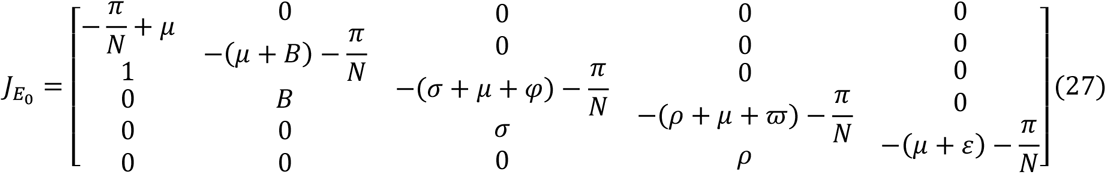

Hence, according to (Gerald, 2012, p255), the Determinant of the Jacobian Matrix 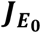 is given by the recursive definition for a 5 × 5 matrix defined as;

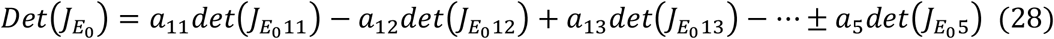

And from (27)

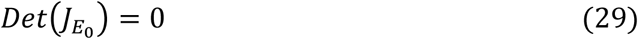

Similarly from the Trace of the Jacobian matrix 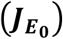 given in equation (27) we have that

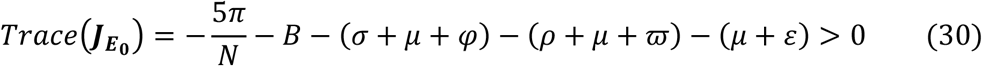

Hence, since 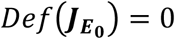 and 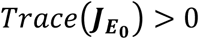 which does not satisfy the prescribed threshold criteria based on Gerald (2012), then the disease free equilibrium (***E***_**0**_) for COVID-19 coronavirus does not satisfy the criteria for a locally or globally asymptotic stability. This implies that as a pandemic as declared by WHO (2020) the COVID-19 coronavirus does not have a curative vaccine yet and precautionary measures are advised through quarantine and observatory procedures.

### 3.4 COMPUTATION OF THE BASIC REPRODUCTIVE NUMBER (*R*_0_) OF THE MODEL

The Basic Reproductive number (*R*_0_) is define as the number of secondary infections that one infectious individual would create over the duration of the infectious period, provided that everyone else is susceptible. *R*_0_ = 1 is a threshold below which the generation of secondary cases is insufficient to maintain the infection in human community. If *R*_0_ < 1, the number of infected individuals will decrease from generation to next and the disease dies out and if *R*_0_ > 1 the number of infected individuals will increase from generation to the next and the disease will persist. To compute the basic reproductive number (*R*_0_) of the model (22) – (26), we employ the next generation method as applied by Diekmann *et. al*., (2009), Van den Driessche and Watmough (2002) and Oduwole and Kimbir (2018).

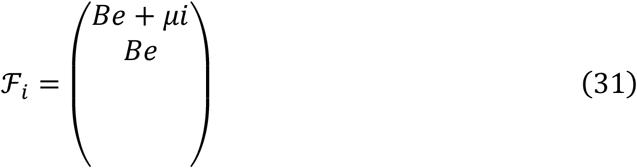

and

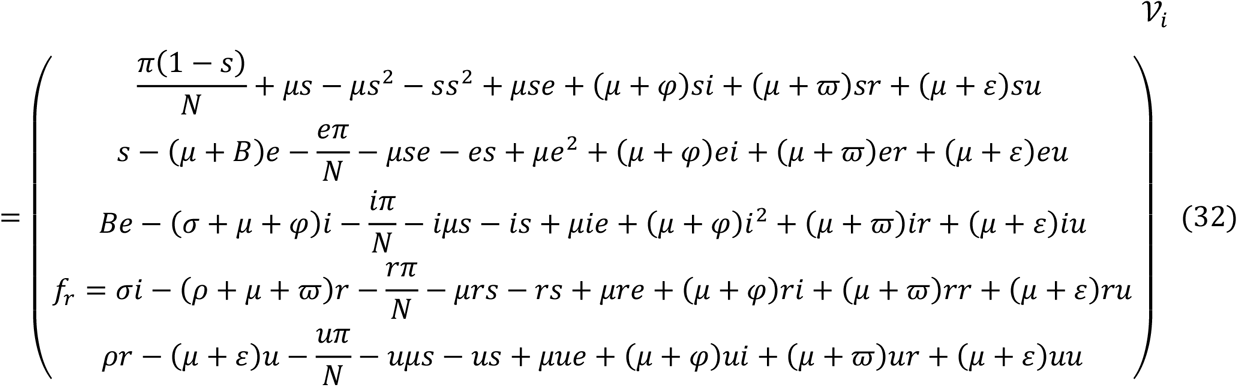

where ℱ _*i*_ and 𝒱_*i*_ are the rate of appearances of new infections in compartment *i* and the transfer of individuals into and out of compartment *i* by all means respectively. Using the linearization method, the associated matrices at disease-fee equilibrium (*E*_0_) and after taking partial derivatives as defined by

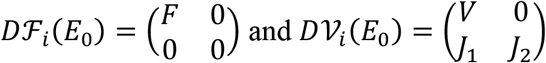

where *F* is nonnegative and *V* is a non-singular matrix, in which both are the *m* × *m* matrices defined by

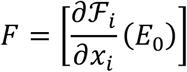

and

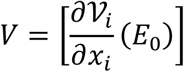

with 1 ≤ *i, j* ≤ *m* and *m* is the number of infected classes.

In particular *m* = 2, we have

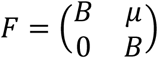

and

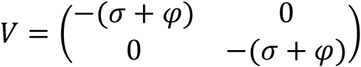

If the inverse of *V* is given as

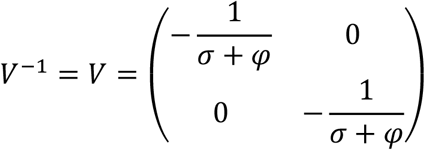

Then the next matrix denoted by *FV*^−1^ is given as

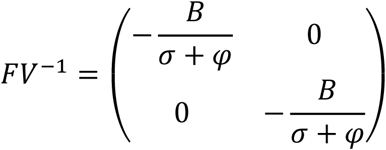

We find the eigenvalues of *FV*^−1^ by setting the determinant |*FV*^−1^ − *γI*| = 0

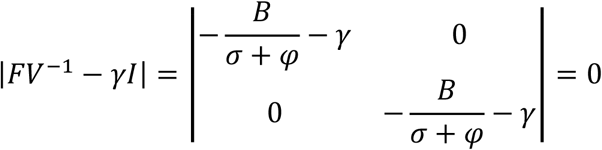

with characteristics polynomial

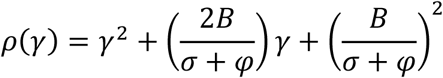

and characteristics equation given as

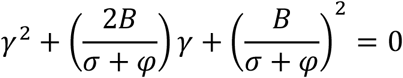

Solving the characteristics equation for the eigenvalues *γ*_1,2_, where *R*_0_ is the maximum of the two eigenvalues *γ*_1,2_. Hence the Basic Reproductive number is the dominant eigenvalues of *FV*^−1^. Thus we have that

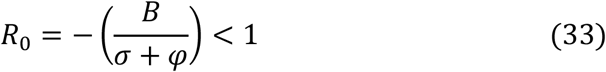

The Basic Reproductive number (*R*_0_ < 1) by Equation (33) shows that there is a chance of decline of secondary infections when the ratio between the incidence rate in the population and the total number of infected population quarantined with observatory procedure. Hence although there currently exist no clinical vaccine for the cure of COVID-19 coronavirus, with Equation (33) there is a high chance of decline in infection when all precautionary measures are observed globally.

## 4. DESCRIPTION AND VALIDATION OF BASELINE PARAMETERS FOR GLOBAL CASES OF COVID-19 CORONAVIRUS

According to the WHO (2020), the total cases of COVID-19 coronavirus worldwide stands: 142,320 and the current total deaths is 5,388 from about 129 countries.

### 4.1 RESULTS AND DISCUSSION

The age-structured deterministic model (11) – (15) was solved numerical using Runge-Kutta-Fehllberg 4-5th order method and implemented using Maple Software. The model equations were first transformed into proportions, thus reducing the model equations to ten differential equations. The parameters used in the implementation of the model are shown in Table 1 below. Parameters were chosen in consonance with the threshold values obtained in the stability analysis of the disease free equilibrium state of the model.

**Table 1:** Estimated values of the parameters used in the Numerical experiments

| Parameters | Values | Data Source | Parameters | Values | Data Source |
| --- | --- | --- | --- | --- | --- |
| $N(0)$ | 7.57bn | WPR (2020) | $\varphi$ | 0.000005* | Assumed |
| $N(1)$ | 149, 292 | WHO (2020) | $\varpi$ | 0.0000007 | JHH (2020) |
| $s(0)$ | 1.0000 | Estimation | $T$ | 14 days | WHO (2020) |
| $e(0)$ | 1.0000 | Estimation | $k$ | 0.5* | Assumed |
| $i(0)$ | 0.00002 | WHO (2020) | $\rho$ | 0.000095 | JHH (2020) |
| $r(0)$ | 0.000095 | JHH (2020) | $\beta$ | 0.00002 | WHO (2020) |
| $u(0)$ | 0.000095 | JHH (2020) | $\sigma$ | 0.28404** | Estimated |
| $\mu$ | 0.000001 | WPR (2020) | $\pi$ | 0.00567* | Assumed |
| $\alpha_0$ | 0.000011 | Batista (2020) | $\varepsilon$ | 0.000095 | JHH (2020) |
| $\alpha$ | | | $B(t)$ | 0.0027** | Assumed |

**Assumed\***: Hypothetical data use for research purpose **Assumed\*\***: Based on Nesteruk (2020).

Hence from Equation (33) the Reproductive Number *R*_0_ = −0.009505 < 1 means there is a 99% chances of secondary infection when an infected population interact by contact with the susceptible population.

Figure 4 clearly shows that over a short period of time the rate of infection of the COVID-19 coronavirus will increase globally before normalizing at a point where future occurrence will be halted due to active quarantine and observatory procedures as prescribed by the WHO (2020) with the hope that a vaccine will be made ready in a shortest future possible.

**Figure 1:**
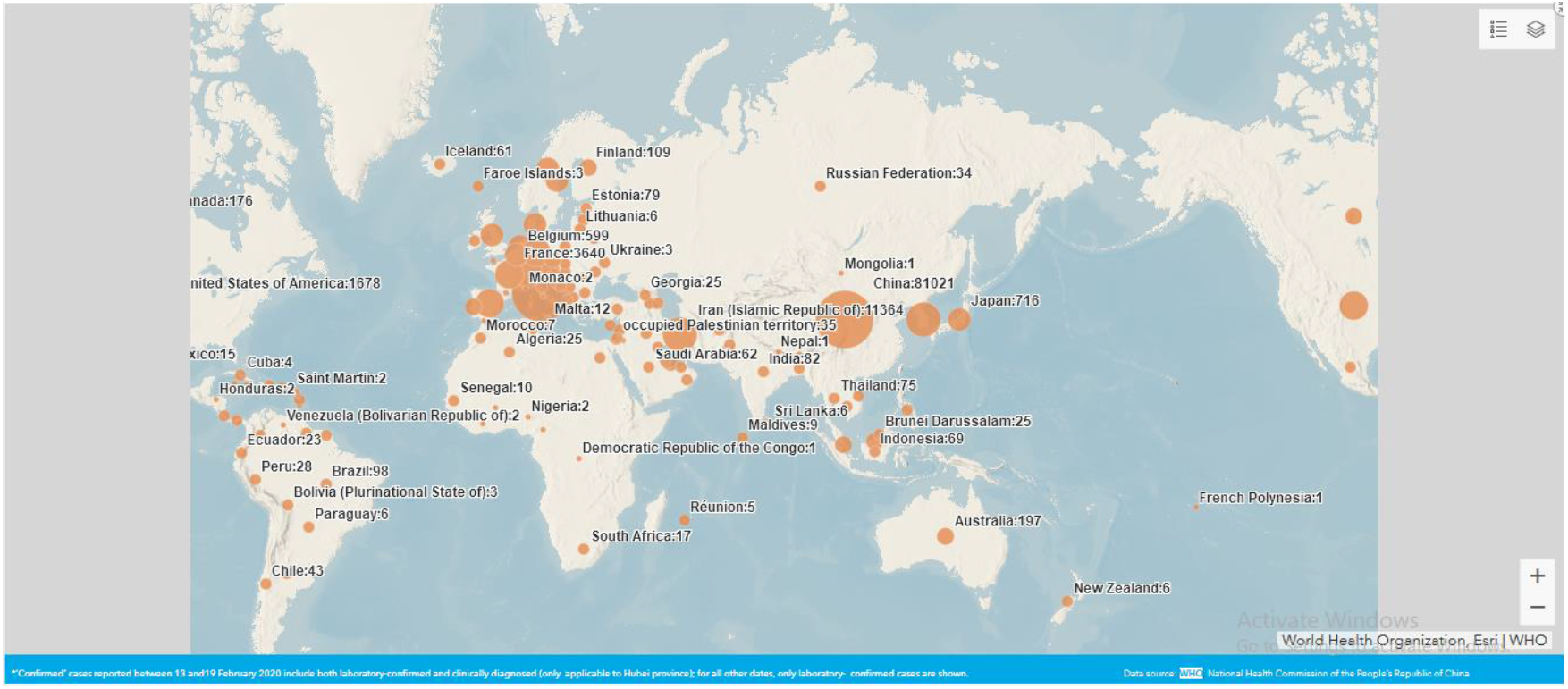
A World Map showing the number of cases per each country with a COVID-19 coronavirus case. (*Source: WHO (2020)*).

**Figure 2:**
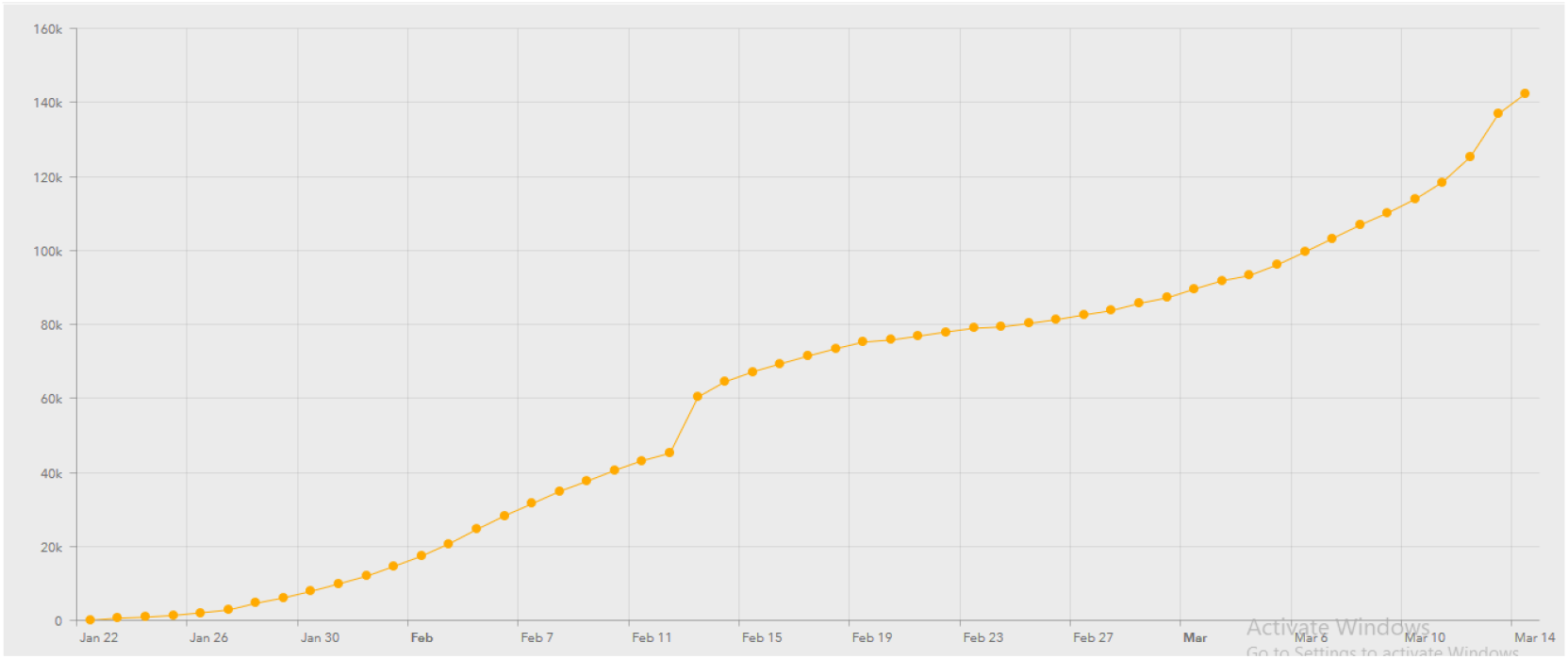
A Cumulative Case chart showing the number of cases with a COVID-19 coronavirus case. (*Source: WHO (2020)*).

**Figure 3:**
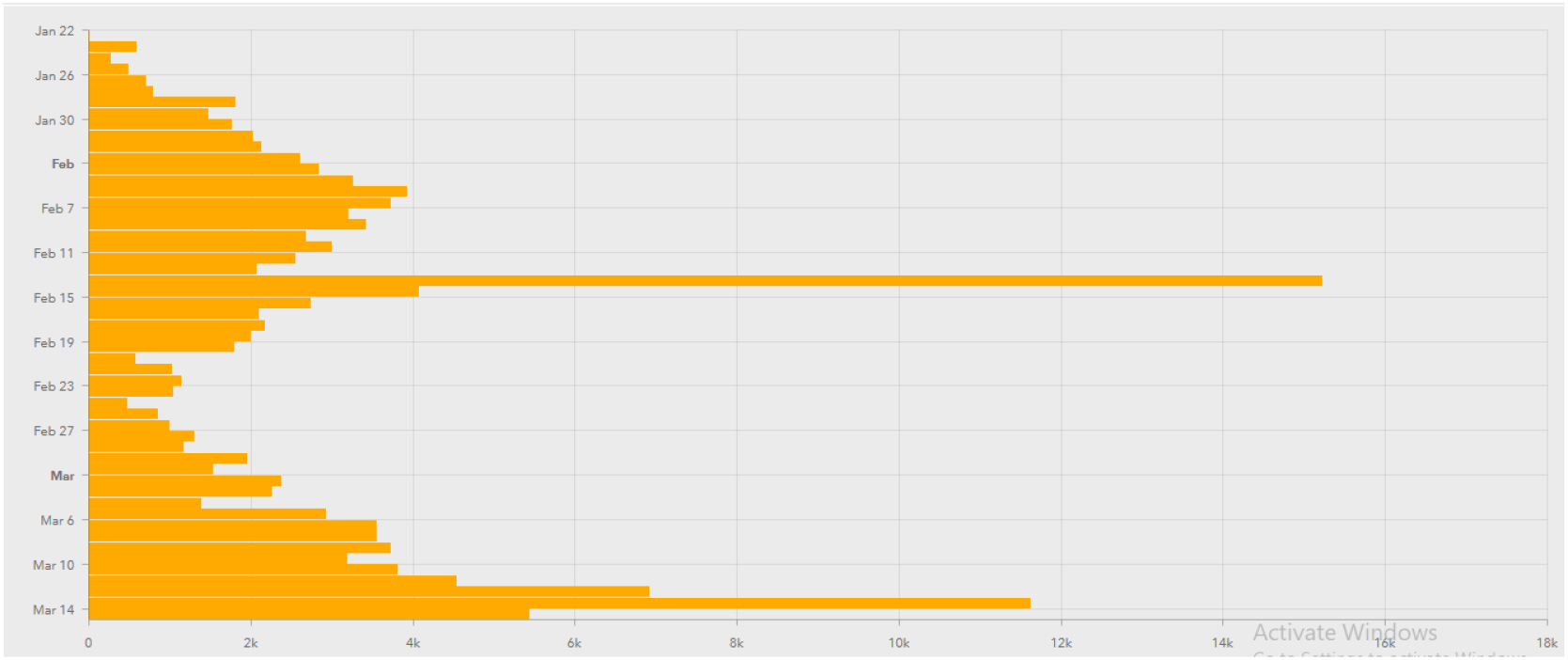
A Case by date of report chart showing the number of cases with a COVID-19 coronavirus case. (*Source: WHO (2020)*).

**Figure 4:**
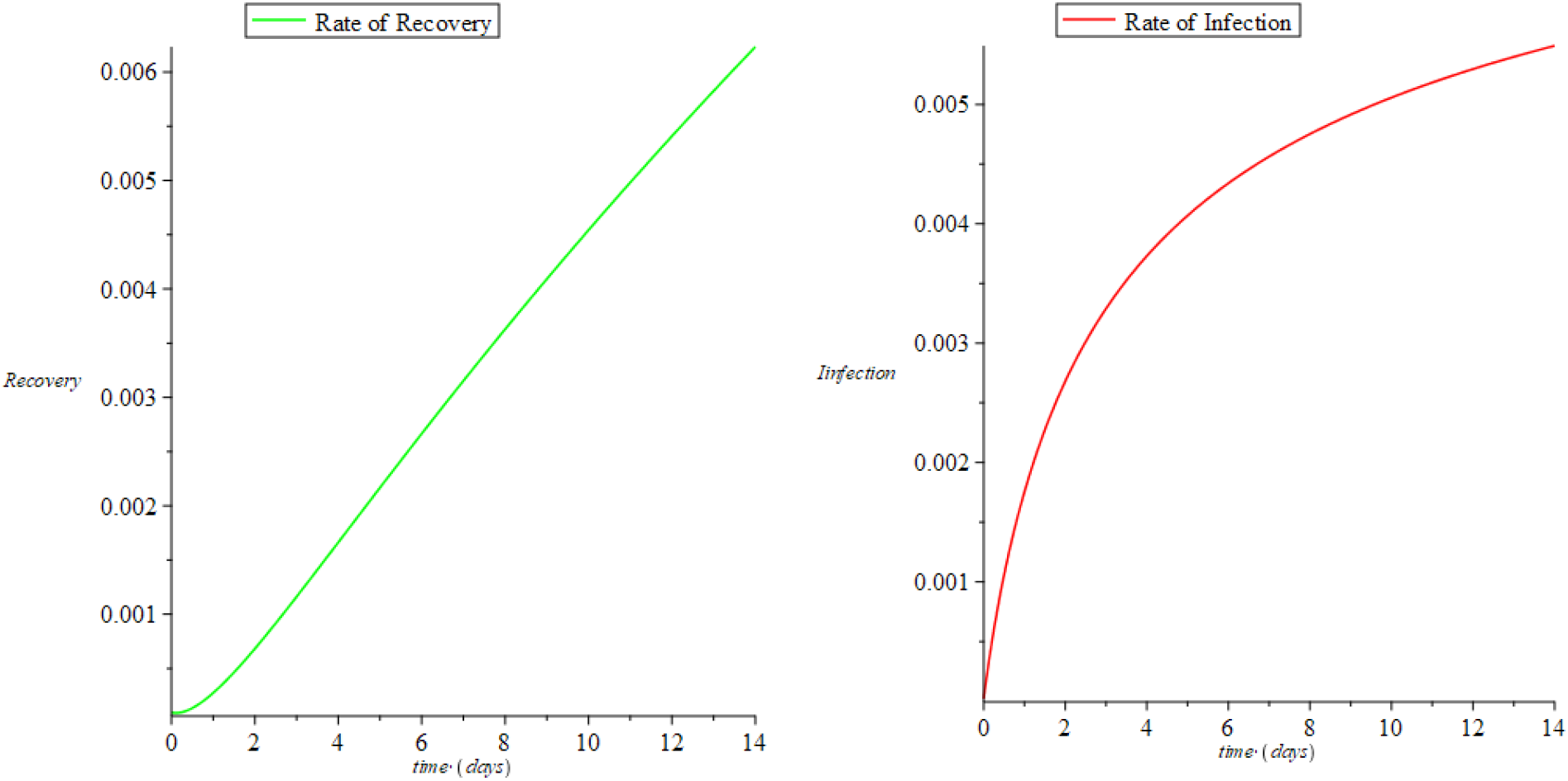
A chart showing the rate of recovery and infection from 1 to 14 days of the global COVID-19 coronavirus pandemic.

Similarly, the chart also reveals that the rate of recovery will continue to increase despite the negligible increase in death rate from the COVID-19 coronavirus globally.

## 5. CONCLUSIONS

In this study, the 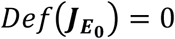 and 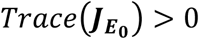 does not satisfy the prescribed threshold criteria based on Gerald (2012), then the disease free equilibrium (***E***_**0**_) for COVID-19 coronavirus does not satisfy the criteria for a locally or globally asymptotic stability. This implies that as a pandemic as declared by WHO (2020) the COVID-19 coronavirus does not have a curative vaccine yet and precautionary measures are advised through quarantine and observatory procedures. Also, the Basic Reproductive number (*R*_0_ < 1) by Equation (33) shows that there is a chance of decline of secondary infections when the ratio between the incidence rate in the population and the total number of infected population quarantined with observatory procedure.

The effort to evaluate the disease equilibrium shows that unless there is a dedicated effort from government, decision makers and stakeholders, the world would hardly be reed of the COVID-19 coronavirus and further spread is eminent. Therefore, as corroborated by the studies of Nesteruk (2020) and Ming and Zhang (2020), the rate of infection will continue to increase despite the increased rate of recovery because of the absence of vaccine at the moment.

Meanwhile, with observatory procedures and quarantine the rate of recovery will not only be enhanced, the chances of total recovery where the virus nor it symptoms will be undetectable is achievable as the rate of detection becomes negligibly small in the bodies of the recovered population.

## Data Availability

The data used in this manuscript were open access data from WHO (2020), JHU (2020), WPR (2020) and Nesteruk (2020) from medrxiv.org

## MAPLE CODE

sol1 := dsolve([diff(N1(t), t) = ((0.00567 + 0.1*10^(-5)*S1(t)*N1(t)) + S1(t)*N1(t)) - 0.1*10^(-5)*E1(t)*N1(t) - 0.6*10^(-5)*I1(t)*N1(t) - 0.17*10^(-5)*R1(t)*N1(t) - 0.000095*U1(t)*S1(t)*N1(t), diff(I1(t), t) = ((0.0027*E1(t) - 0.284046*I1(t) - 0.00567*I1(t)/N1(t) - 0.1*10^(- 5)*I1(t)*S1(t)) - I1(t)*S1(t)) + 0.1*10^(-5)*I1(t)*E1(t) + (0.6*10^(-5)*I1(t))*I1(t) + 0.17*10^(-5)*I1(t)*R1(t) + 0.000095*I1(t)*U1(t), diff(E1(t), t) = ((S1(t) - 0.002701*E1(t) - 0.00567*E1(t)/N1(t) - 0.1*10^(-5)*S1(t)*E1(t)) - S1(t)*E1(t)) + (0.1*10^(-5)*E1(t))*E1(t) + 0.6*10^(- 5)*I1(t)*E1(t) + 0.17*10^(-5)*E1(t)*R1(t) + 0.000095*E1(t)*U1(t), diff(R1(t), t) = ((0.28404*I1(t) - 0.0000967*R1(t) - 0.00567*R1(t)/N1(t) - 0.1*10^(-5)*R1(t)*S1(t)) - R1(t)*S1(t)) + 0.1*10^(-5)*E1(t)*R1(t) + 0.6*10^(-5)*I1(t)*R1(t) + (0.17*10^(-5)*R1(t))*R1(t) + 0.000095*R1(t)*U1(t), diff(U1(t), t) = ((0.000095*R1(t) - 0.000096*U1(t) - 0.000096*U1(t)/N1(t) - 0.1*10^(-5)*U1(t)*S1(t)) - U1(t)*S1(t)) + 0.1*10^(-5)*E1(t)*U1(t) + 0.6*10^(-5)*I1(t)*U1(t) + 0.17*10^(-5)*R1(t)*U1(t) + (0.000095*U1(t))*U1(t), diff(S1(t), t) = 0.00567*(1 - S1(t))/N1(t) + 0.1*10^(-5)*S1(t) - (0.1*10^(-5)*S1(t))*S1(t) - S1(t)*S1(t)*S1(t) + 0.1*10^(-5)*S1(t)*E1(t) + 0.6*10^(-5)*I1(t)*S1(t) + 0.17*10^(- 5)*R1(t)*S1(t) + 0.000095*U1(t)*S1(t), N1(0) = 7570000000, I1(0) = 0.00002, E1(0) = 1.00000, R1(0) = 0.000095, U1(0) = 0.000095, S1(0) = 1.00000], numeric);

plots[odeplot](sol1, [[t, R1(t), color = red], [t, U1(t), color = blue], [t, I1(t), color = green]], 0 ‥ 14);

